# The TB vaccine clinical trial centre directory: an inventory of clinical trial centres in sub-Saharan Africa

**DOI:** 10.1101/2023.10.04.23296539

**Authors:** Puck T. Pelzer, Marit Holleman, Michelle E.H. Helinski, Ana Lucia Weinberg, Pauline Beattie, Thomas Nyirenda, Job van Rest, Gerald Voss

## Abstract

**Background:** There are over ten vaccine candidates for tuberculosis (TB) in the clinical pipeline that require testing in TB-prevalent populations. To accelerate the clinical development of TB vaccines, a directory of clinical trial centres was established in sub-Saharan Africa (SSA) to assess capacity for conducting late-stage TB vaccine trials.

**Methods:** TB vaccine-related parameters were identified, and trial centres in SSA were identified and prioritized based on whether they had experience with TB or non-TB vaccine trials. A survey was sent to identified centres, and the resulting directory presents their capacity for TB vaccine trials. Centres that deemed themselves eligible for TB vaccine trials also had the option to contribute their information to the survey. This article provides an overview of the TB vaccine clinical trial centre directory, including the number and distribution of centres, their general characteristics, and their experience with prior TB vaccine trials. It includes information on the capacity of the centres, such as laboratory biosafety level, patient support, and community engagement. It also includes a case study to demonstrate how the directory can be used to identify trial centres with specific capabilities needed for a particular TB vaccine trial.

**Results:** Of the 134 identified centres, 56 responded by providing information. Of these centres, 51 (91%) had phase 3 clinical trial experience and previous TB trials were conducted at 38 centres. Regarding TB vaccine trials, 19 centres conducted prevention of disease trials, 14 conducted prevention of infection trials, and 27 had no experience with TB vaccine clinical trials. From the respondents, 29 centers in South Africa were identified that could potentially conduct TB vaccine trials, followed by Tanzania (5), Kenya (5), Nigeria (3), and Uganda and Ethiopia (2 each). Trial sites in other countries were underrepresented, based on this survey.

**Conclusion:** The establishment of a clinical trial centre directory can provide a basis for decision-making by various stakeholders. Despite some limitations in survey methodology, the findings suggest opportunities for expanding the evaluation of clinical trial capacity in other disease-prevalent countries and continents. Such data would be valuable in further enriching the Clinical Trial Community which a resource that geographically highlights clinical trial investments and capacities in African research ecosystem.

**Summary Points:**

- New TB vaccine candidates need to be assessed in clinical trials in countries with high rates of TB in the coming years.
- An open-access directory of TB vaccine clinical trial centres in sub-Saharan Africa was established, providing an overview of the capacity to conduct clinical trials for TB vaccine candidates (http://www.edctp.org/our-work/coordination-tb-vaccine-funded-research/directory-tb-vaccine-clinical-trial-sites-sub-saharan-africa/).
- The directory is intended for clinical triallists, funders, policymakers, and researchers to accelerate the clinical development of novel TB vaccines by providing useful information.
- Regular updates are necessary to ensure the directory remains relevant for vaccine development and feeds into the continental Clinical Trials Community (https://ctc.africa/).

## Introduction

Tuberculosis (TB) remains a significant global health threat, responsible for approximately 1.3 million deaths in 2020 alone (1). Immunization against TB is a critical component of the fight against this disease, yet the only registered vaccine currently available is the bacillus Calmette Guerin vaccine (BCG) (2). Although the BCG vaccine has been shown to prevent certain types of TB (i.e., meningitis and disseminated TB in young children), its protection against pulmonary TB is variable (3,4). Currently BCG is only recommended for new-borns and is contra-indicated in HIV-positive populations. Given the ongoing threat of TB and the ambitious “End TB” goals set out by the World Health Organization, the development of new and improved vaccines is urgently needed to accelerate progress towards elimination.

The current pipeline includes vaccines based on recombinant viral vectors, live attenuated *Mycobacterium tuberculosis*, improved BCGs, inactivated whole cells and adjuvanted proteins (5). At present, there are five vaccine candidates in phase 3 clinical trials, and two additional candidates will enter phase 3 soon. VPM1002 targets prevention of infection (PoI), prevention of disease (PoD) and prevention of recurrence (PoR) in different phase 3 trials. MIP/Immuvac, GamTBvac and MTBVAC are aiming at PoD. The M72/AS01E candidate is scheduled for a phase 3 efficacy trial for PoD and PoI. BCG revaccination will be evaluated in a Phase 3 trial for PoI (6).

In the recent past, few TB vaccine trials have been conducted in the world. The capacity needed for TB vaccine trials differs from the requirements of trials for TB diagnostics, drugs, and preventive treatment. Whereas vaccine trials are conducted in healthy populations or infected populations, trials for drugs and diagnostics require TB patients and trials for preventive treatment require populations with high TB infection rates or substantial TB-HIV co-infection prevalence. For TB vaccine clinical trials, follow-up periods typically range from several months to several years, depending on the specific vaccine candidate and study design. For TB diagnostic tests, follow-up periods in clinical trials can vary widely based on the type of diagnostic being evaluated. They may range from a few weeks to several months. Furthermore, the number of participants needed for TB vaccine trials is substantially higher. To cover the host and bacteriological genetic variety, multiple locations are required for phase 3 TB vaccine trials to allow the licensing of new vaccines in different countries. In addition, trial centres require diverse and specific capacity and capabilities for the different targeted endpoints. For example, PoD trials require a substantially larger sample size with prolonged follow-up, as compared to PoI trials.

To further advance TB vaccine development, the vaccine candidates in late-stage development need to be tested in TB prevalent populations over the coming years. Performing such work requires access to clinical trial centres with adequate capacity around the world, particularly for phase 3 licensure trials. Developing a searchable directory of clinical trial centres and their capacity is expected to aid in the advancement of the development of TB vaccines. To this end, we have created a directory of clinical centres in sub-Saharan Africa (SSA) suitable for future TB vaccine studies.

## Methods

To gather information from as many trial centres in sub-Saharan Africa (SSA) as possible, a multistep approach was employed, which included identifying key parameters, selecting centres for outreach, designing a survey, collecting, and managing data. A desk review was conducted to identify the essential parameters for the TB vaccine trials and the trial centre directory, which included a review of the existing literature on TB vaccine trials and reports. Then, four experts were invited to review the parameters and data collection tool, based on their expertise in the design of TB vaccine trials, TB in general, and survey design.

The resulting survey and directory parameters consisted of the centre characteristics, the type of disease targets and intervention, and expertise and the capacity (Table 1). A clinical trial centre directory template was developed along with data collection tools, including an Excel file and online survey. Data collection was carried out using a Microsoft Forms online survey tool, which contained 40 questions in English (Supplement 1).

**Table 1.**
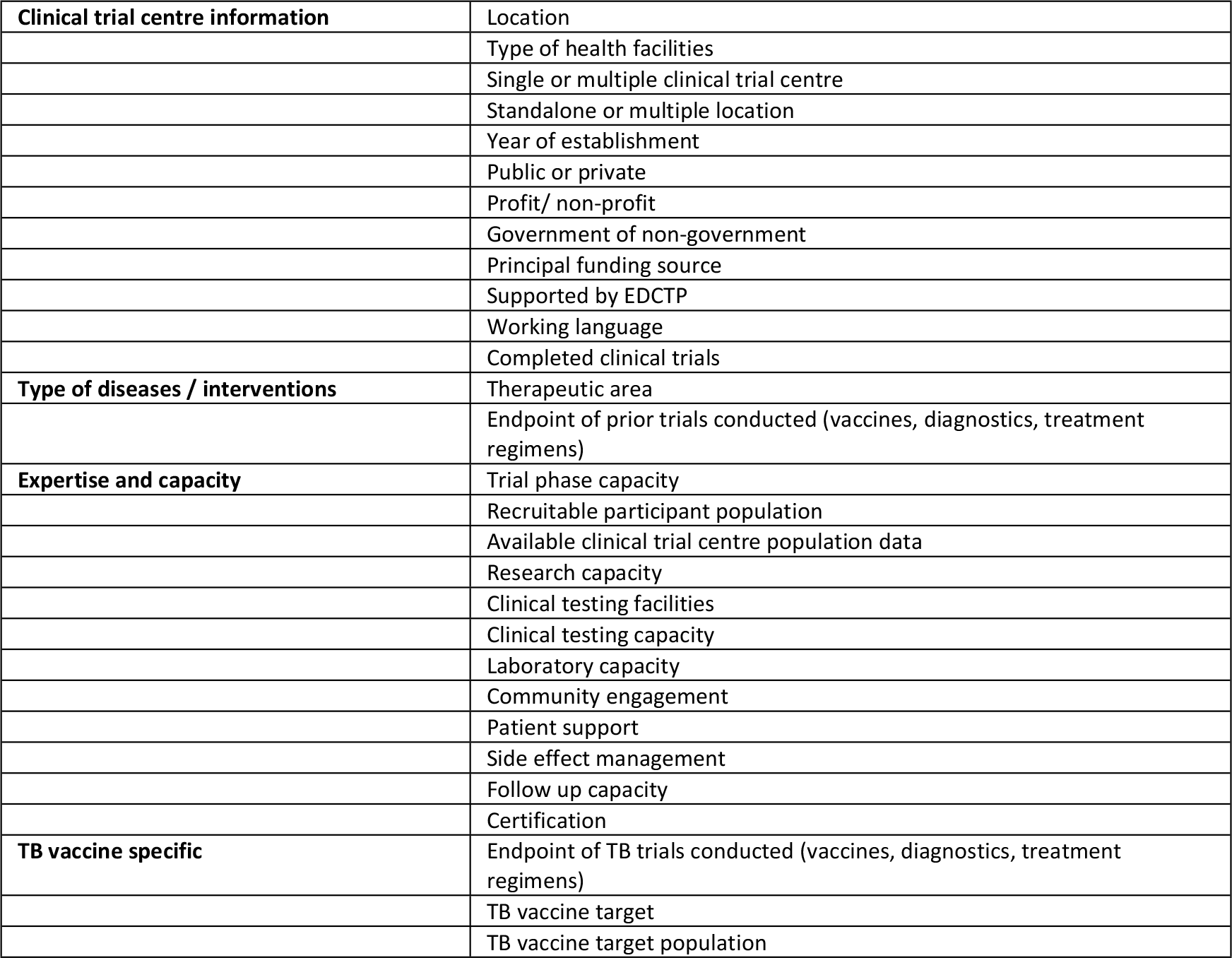
Parameters considered in the TB vaccine clinical trial centre directory: centre characteristics, type of disease targets, intervention expertise and capacity, TB vaccine.

To identify clinical trial centres for outreach, a desk review of SSA centres was conducted using various websites, including ClinicalTrials.gov, Centerfinder, Global Health Trials, Centrewatch, AAS trial centres, and AGT network. Each centre was evaluated for suitability for the TB vaccine trial centre directory, with shortlisting based on prior experience with TB vaccine trials or trials related to TB and non-TB vaccine trials. In addition, centres were included based on expert recommendation, and contact persons were identified through online searches and referrals from the field. Centres that considered themselves eligible for conducting TB vaccine trials were also included.

The respective contact persons for the shortlisted centres were invited by email to complete the online survey, with survey responses collected between April and August 2020. Centres that did not respond received up to two reminders, and additional contact details were sought to solicit a response. The first round of data collection closed in 2020, and in 2022, the survey was reopened to include clinical trial centres that considered themselves eligible or wished to be included in the directory.

The survey data were cleaned using Stata v15.1 (Stata Corp, College Station, TX, USA). Data from each centre were manually inspected, duplicates were removed, and data entry errors were corrected. The respondents were asked to validate the data if necessary. The clinical trial centre directory entries were summarized by general centre characteristics and TB vaccine trial-specific characteristics. For each, the potential trial endpoints, recruitable population, and phase of clinical trials were highlighted, along with TB incidence in the country as reported by WHO (8) and population and country size from World Bank estimates (9) for all countries in SSA and for included trial centres.

## Results

The survey data were compiled into the TB vaccine clinical trial centre directory. The directory is publicly accessible on the EDCTP website. Figure 1 shows the website of the trial centre directory. Users can review information on clinical trial centres in each country by clicking on an interactive map to review key details for that location. In addition, the entire dataset including additional parameters and the data dictionary are available as downloads from the website.

**Figure 1.**
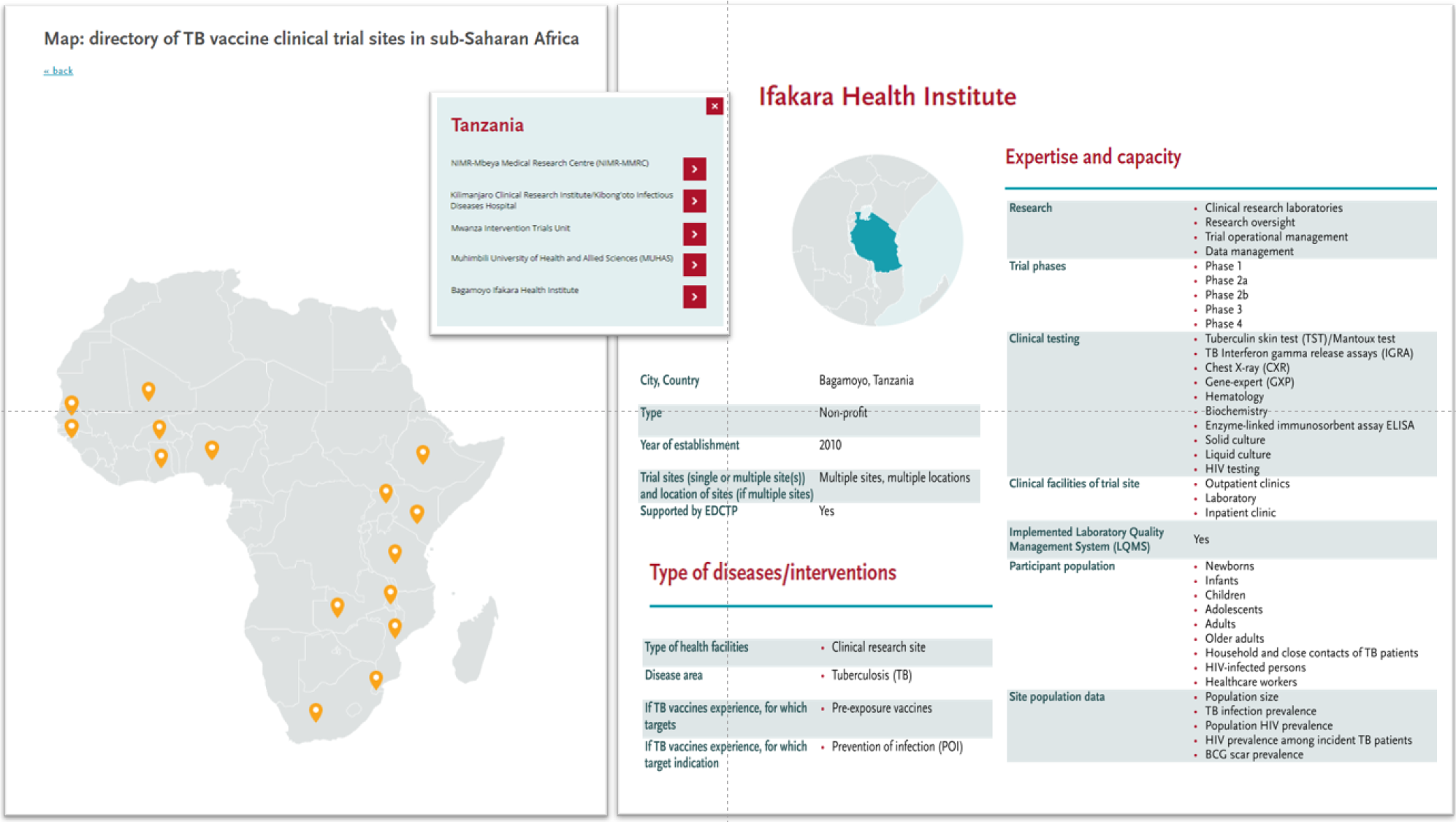
Website of the TB vaccine trial centre directory, and an example of key trial centre information provided online.

### General characteristics

Our study shortlisted 134 clinical trial centres in SSA, of which 43% responded to our survey (Supplement 2) and were included in the trial centre directory. The first version of the directory, which included data from 45 centres, was established in May 2021. Between May 2021 and June 2022, an additional 11 centres were included based on self-assessment. The directory was last updated in June 2022.

The majority of the 56 centres in the directory are located in South Africa (29), followed by Tanzania and Kenya with five centres each, and three in Nigeria. Two centres are in Uganda and Ethiopia, and the remaining nine countries have one centre each (Table 3). Out of the 56 centres, 22 were standalone clinical trial centres and 33 were clinical trial centres with multiple sites. The year of establishment of these centres ranges between 1934 and 2021. Most of the centres (46) were non-profit organisations. On average, the assessed centres had completed 26 clinical trials (TB or non-TB) and had follow-up capacity of 10 years or more at 20 centres, 5 years at 21 centres, and less than 3 years at 13 centres (Figure 2, Panel A). Patient support, community engagement, and side effect management were reported to be in place at most of the centres (Figure 2, Panel B). About one-third of the centres (24) reported having laboratory biosafety level (BSL) 3 capabilities, and four had BSL4 capabilities at the centre or affiliated laboratory (Figure 2, Panel C). Most centres were clinical research centres (51), while 19 were academic institutions (Figure 2, Panel D). Thirty-eight centres had a clinical research focus on TB (Figure 2, Panel E).

**Figure 2:**
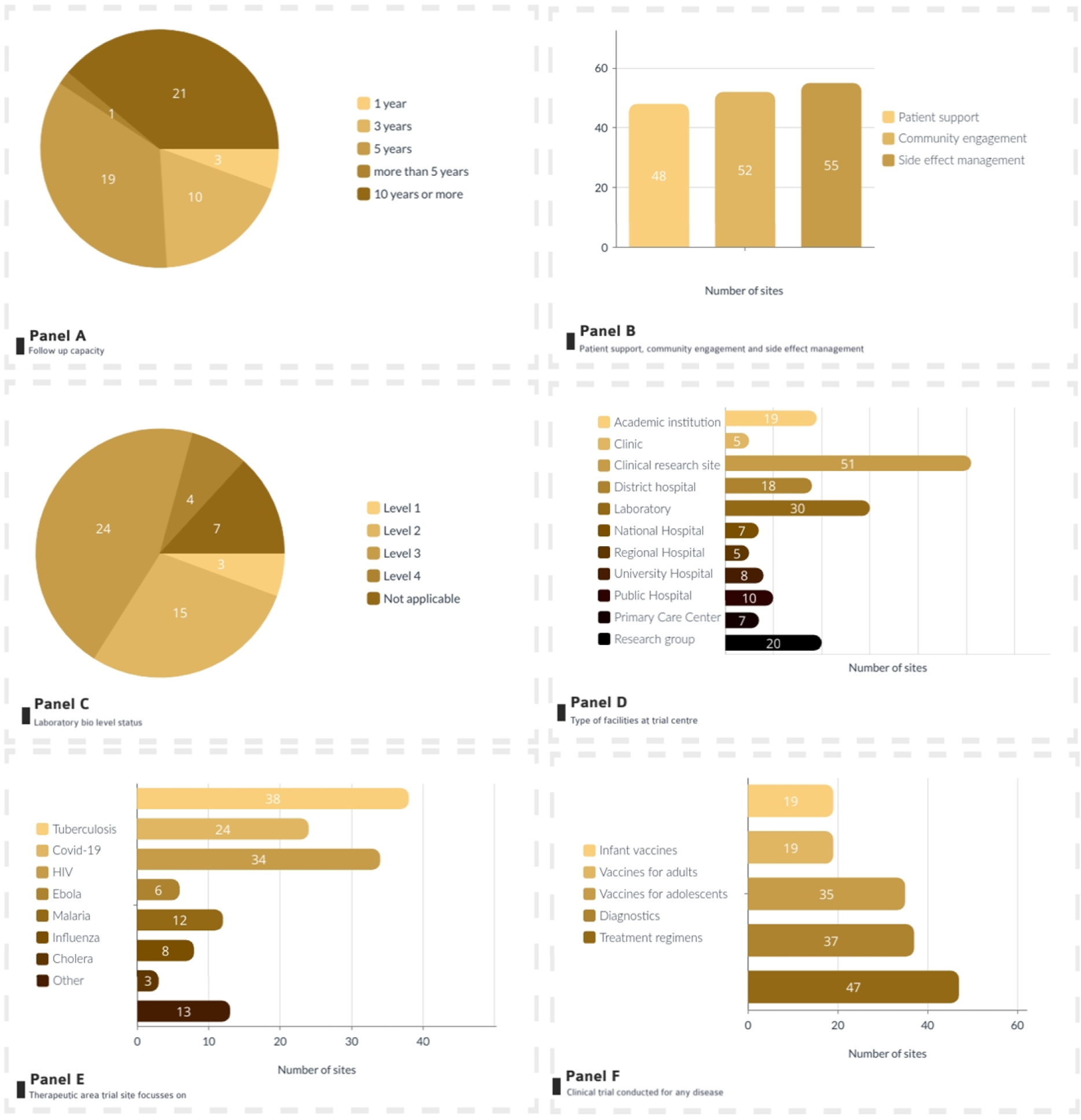
Overview of centre characteristics overview of trial centres included in the directory. Panel A: The follow-up capacity of the trial centre | Panel B: patient support, community engagement and side effect management | Panel C Laboratory biosafety level | Panel D: Type of facilities at trial centre| Panel E: Therapeutic area trial centre focusses on | Panel F: Clinical trials conducted for any disease. Panel B,D,E, F are not mutually exclusive

Twenty-seven clinical trial centres reported prior TB vaccine l trial experience, with the majority of these having experience in TB vaccine trials in adult populations. (Figure 3, Panel A). Twenty centres had capacity for preexposure TB vaccines and 17 for post (Figure 3, Panel B). Most of the 27 centres had experience with PoD (19) and/or PoI (14) trial endpoint capacity (Figure 3, Panel E). Tuberculin skin testing was available at 46 centres, interferon gamma release assay at 32 centres, culture testing at 30 centres and HIV testing at 53 centres (Figure 3, Panel D).

**Figure 3:**
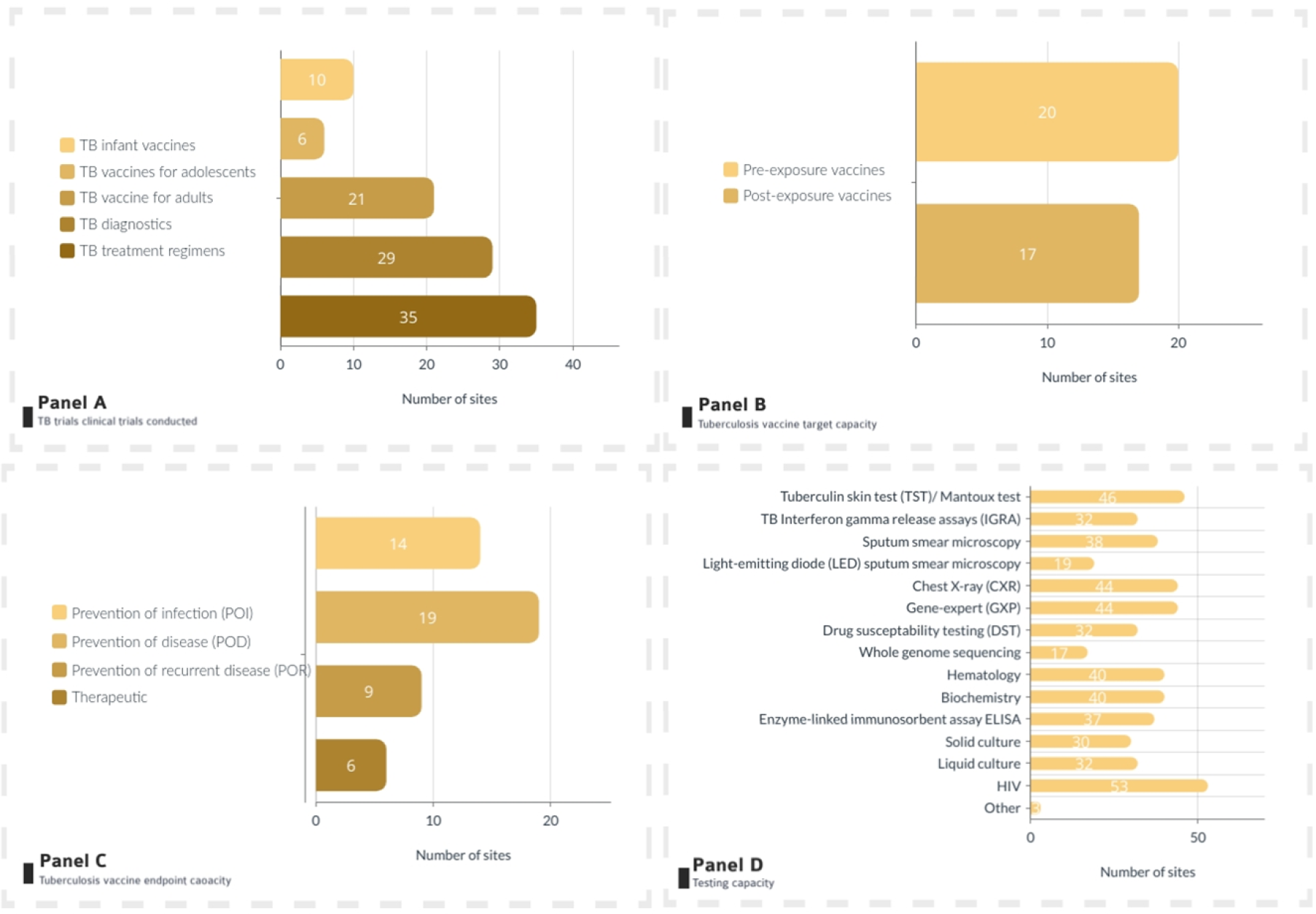
Overview of TB clinical trial specific characteristics overview of trial centres included in the directory. Panel A: TB trials clinical trials conducted | Panel B: Tuberculosis vaccines target capacity | Panel C: Tuberculosis vaccines endpoint | Panel D: Testing capacity. Panel A-D are not mutually exclusive.

From the respondents, South Africa had the most centres with experience in TB vaccine trials (16 centres), followed by Tanzania (3 centres), Kenya (2 centres) and Ethiopia, Guinea-Bissau, Mozambique, Senegal, Uganda and Zambia with one centre each (Table 2). PoD trial endpoint capacity was available in 19 centres, of which the majority was in South Africa. The same was observed for PoI trial endpoint capacity, with 10 out of 14 centres being located in South Africa. PoR capacity was mostly also seen in South Africa (7 out of 9 centres). Of all the trial centres, 51 had phase 3 capacity (any disease). In terms of recruitable populations, centres had experience in working with adult populations. Less experience was among adolescents (46/56) and least experience with neonates (26/56).

**Table 2.** Clinical trial centre capacity per country.

| Country | Trial centres | Experience with TB vaccine trials |  |  | Experience with trial phases (any disease/intervention) |  |  |  |  | Recruitable population |  |  |  |
| --- | --- | --- | --- | --- | --- | --- | --- | --- | --- | --- | --- | --- | --- |
|  |  | PoD | PoI | PoR | I | II a | II b | III | IV | Adults | Adolescents | Children | Neonates |
| Burkina Faso | 1 | 0 | 0 | 0 | 0 | 1 | 1 | 1 | 1 | 1 | 1 | 1 | 1 |
| Cameroon | 1 | 0 | 0 | 0 | 0 | 0 | 1 | 1 | 1 | 1 | 1 | 0 | 0 |
| Eswatini | 1 | 0 | 0 | 0 | 0 | 1 | 1 | 0 | 0 | 1 | 1 | 1 | 1 |
| Ethiopia | 2 | 1 | 0 | 0 | 2 | 2 | 2 | 2 | 1 | 2 | 2 | 0 | 0 |
| Ghana | 1 | 0 | 0 | 0 | 0 | 1 | 1 | 1 | 1 | 1 | 1 | 1 | 1 |
| *Guinea-Bissau* | 1 | 0 | 0 | 0 | 0 | 0 | 1 | 1 | 1 | 1 | 1 | 1 | 1 |
| *Kenya* | 5 | 1 | 2 | 1 | 4 | 4 | 3 | 4 | 4 | 5 | 5 | 3 | 3 |
| *Malawi* | 1 | 0 | 0 | 0 | 0 | 0 | 0 | 1 | 1 | 1 | 1 | 1 | 1 |
| *Mali* | 1 | 0 | 0 | 0 | 1 | 1 | 0 | 0 | 0 | 1 | 1 | 1 | 0 |
| *Mozambique* | 1 | 1 | 0 | 0 | 0 | 1 | 1 | 1 | 1 | 1 | 1 | 1 | 1 |
| *Nigeria* | 3 | 0 | 0 | 0 | 1 | 1 | 2 | 3 | 2 | 3 | 2 | 1 | 1 |
| *Senegal* | 1 | 0 | 0 | 0 | 1 | 1 | 1 | 1 | 0 | 1 | 1 | 1 | 1 |
| *South Africa* | 29 | 14 | 10 | 7 | 13 | 21 | 27 | 27 | 16 | 29 | 21 | 13 | 10 |
| *Tanzania* | 5 | 1 | 2 | 1 | 3 | 3 | 5 | 5 | 4 | 5 | 4 | 4 | 4 |
| *Uganda* | 2 | 0 | 0 | 0 | 1 | 1 | 1 | 2 | 1 | 2 | 2 | 1 | 1 |
| *Zambia* | 1 | 1 | 0 | 0 | 0 | 0 | 1 | 1 | 1 | 1 | 1 | 0 | 0 |
| **Total** | **56** | **19** | **14** | **9** | **26** | **38** | **48** | **51** | **35** | **56** | **46** | **30** | **26** |

PoD = prevention of disease, Pol= prevention of infection, PoR = prevention of recurrence

Figure 4 shows an overview of WHO TB incidence rates per 100,000 in SSA, highlighting potential TB vaccine clinical trial centres included in the directory. The six countries with the highest relative incidence rate per 100,000 in SSA either had no trial centre identified (Republic of Congo, Lesotho, Angola, Central African Republic, Namibia), or had one centre identified that did not respond (Gabon). Of the included trial centres, South Africa has the highest incidence relative to the population and also the largest number of clinical trial centres. The four subsequent countries with the highest incidence were Mozambique, Guinea-Bissau, Zambia and Eswatini (Supplement 2). Each of these countries has one trial centre that responded to the survey, potentially suitable for TB vaccine trials in the country. It should be noted here that Eswatini and Guinea-Bissau are relatively small in terms of population size, and for Zambia five additional centres were identified, but these did not respond to the survey (Supplement 2).

**Figure 4.**
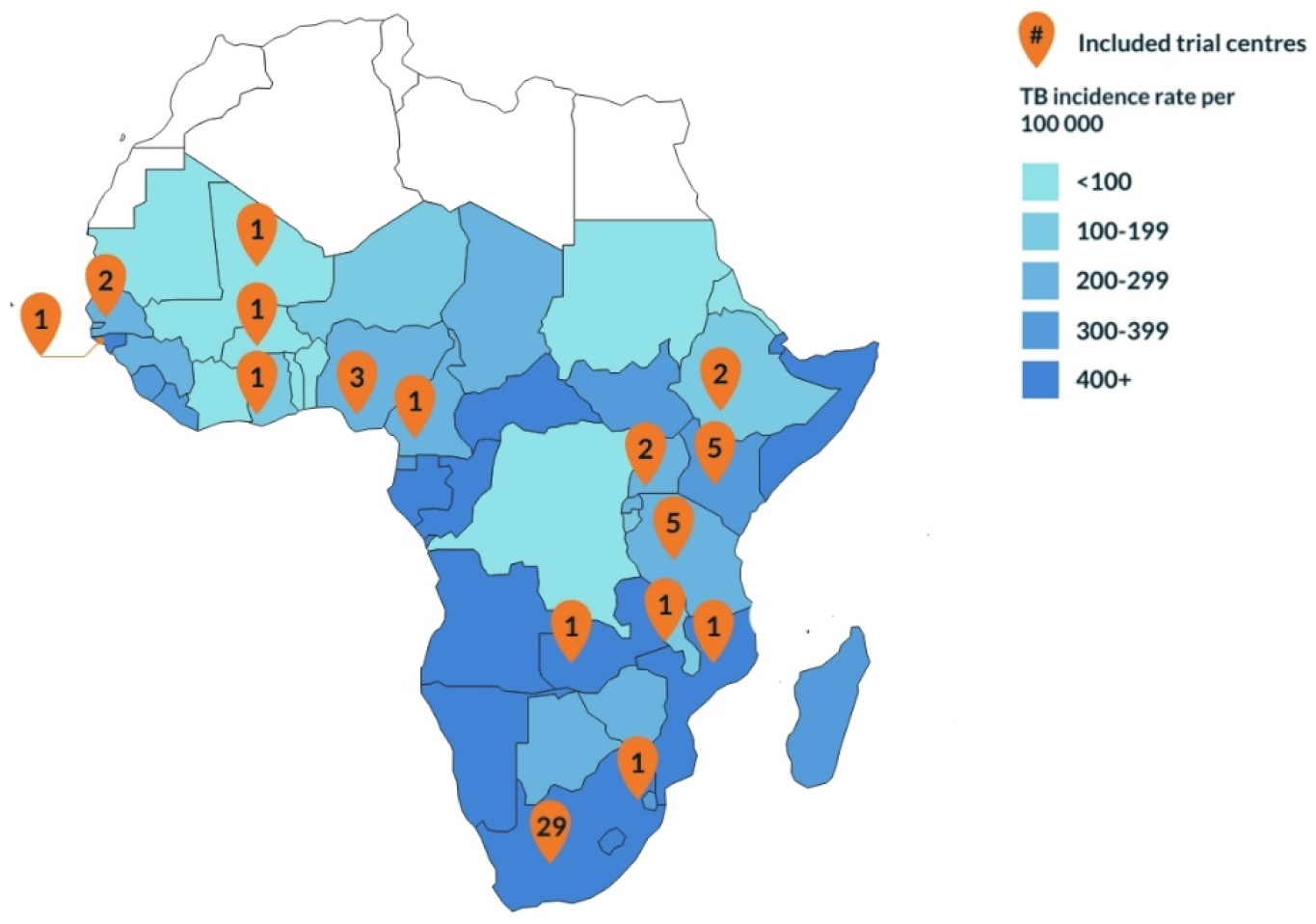
Map of TB vaccine trial centres included in the directory and WHO TB incidence rates per 100 000 population.

One of the applications of the directory is to identify trial centres that have a specific combination of capabilities needed for a particular TB vaccine trial. Figure 5 shows the applicability of the trial centre directory for a case study that would require phase 3 trial capabilities, with experience in the PoD trial endpoint among adolescent in a WHO high TB burden country, where infection testing would be carried out through IGRA and GeneXpert for TB. Participants would be approached through an in-patient clinic. Applying these search criteria in the clinical trial centre directory resulted in five potential centres that could be used, as shown in Figure 5.

**Figure 5.**
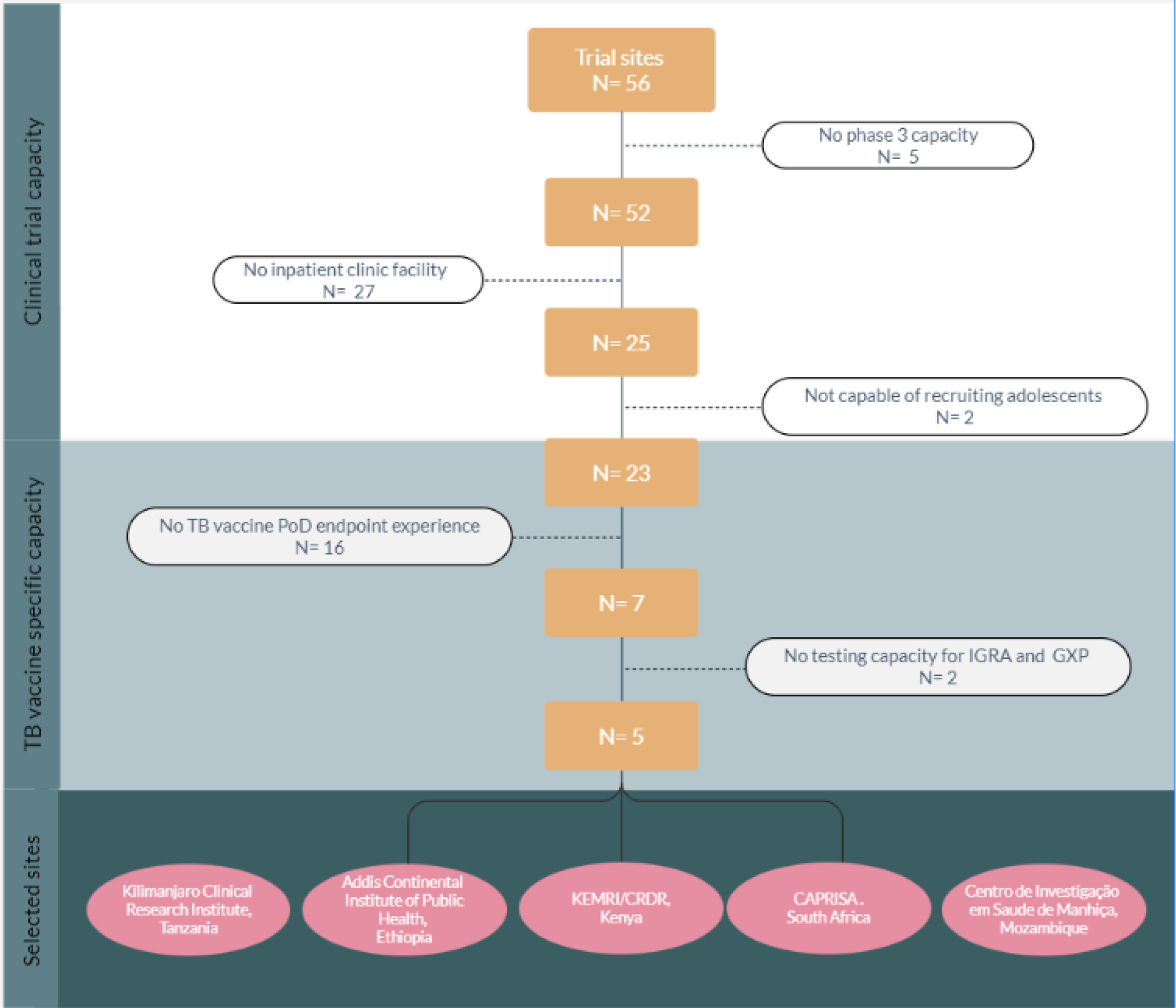
Case study using the TB vaccine clinical trial centre directory.

## Discussion

This project established a directory of clinical trial centres capable of conducting TB vaccine trials in SSA. The evaluation of the compiled trial centre directory highlights the need to develop more trial centres across the four regions of SSA reducing reliance on the South African sites. This is especially relevant considering a pipeline of promising vaccine candidates that are being prepared for phase 3 studies (7). In terms of target populations, the evaluated centres had sufficient capacity to recruit adults, but the limitations on neonates was most striking. Therefore, it is necessary to strengthen the capacity of the trial centres to conduct TB vaccine trials for different indications.

The clinical trial centre assessment had some limitations. In the directory, only responding sites were included, while in some counties additional trial centres exist, making it difficult to have a comprehensive overview for any such given country. There is over-representation of South African centres, which could be attributed to the country’s investments in TB vaccine centres. In addition, the information provided was self-assessment by the centres and not further validated. Just under half of the centres contacted responded (43%). Possible reasons affecting the response rate could have been language barriers, timing of survey during the COVID-19 pandemic when research centres were prioritising response to COVID, difficulties identifying the most appropriate contact person for the centre and staff turnover. Future efforts in this area should aim to include centres and countries that were missed as well as an expansion to other geographical (global) areas including translating the survey in other languages e.g., French and Portuguese. This will be a necessary effort, as TB vaccine trials will be conducted in countries outside of Africa where TB is endemic.

To keep the TB vaccine clinical trial centre directory relevant and useful, regular updates are needed. In terms of epidemiological indicators, this mapping assessment was limited to national data. Ideally, centres should be evaluated by their own age-specific epidemiological indicators (7). The latest estimates on TB infection (8) are from 2014, making them somewhat outdated. In terms of centre capacity, including diagnostics, this may evolve over time and to remain relevant, centres should be regularly contacted to update information.

Despite the limitations described above the directory allows to identify the registered centres that have experience and capacity in terms of TB vaccines, complementing other initiatives to map clinical trial centers such as the African clinical trials community (https://www.ctc.africa/about), which is a database of clinical trials investments and clinical sites including their capacities for all diseases. The study did not investigate the factors that influence the establishment of TB vaccine trial centres in individual countries. Thus, further work should also investigate what incentivises institutions to become a TB vaccine trial centre. For example, prior evaluations of TB vaccine trial centres indicated that difficulties with logistics and operations could pose a problem in terms of trial execution (9,10). Kaguthi et al. (11) discuss lessons learned from development of a TB vaccine trial centre for design and implementation of trials in Africa. They highlight that performing exhaustive epidemiological research to offer background incidence and prevalence rates of TB for clinical trials, and the inclusion of investigators in discussions about tests to be run on bio-banked samples are some of the key lessons learned in centre development.

Our study focused on SSA to demonstrate efficacy under varying complex epidemiological and clinical conditions, but for the global development of TB vaccines, other countries and populations must be included in late-stage TB vaccine trials. In a recent study (12), experts indicated that nationally conducted trials could be a requirement in the country prior to implementation, indicating the importance of centre development throughout the globe and especially in TB endemic areas in order to ensure adequate human and mycobacterial diversity. Therefore, the expansion of the clinical trial centre directory for TB vaccines to other geographies may prove useful.

## Conclusions

A survey of centres in TB endemic countries through an online questionnaire resulted in the establishment of a TB vaccine clinical trial centre directory providing information that may be helpful to accelerate the clinical development of TB vaccines. The directory is intended as a tool for clinical trialists, vaccine developers, funders, policymakers, and researchers. It provides a snapshot in time and needs to be updated periodically to remain relevant. The capacity gaps exposed by the directory showed that there is a need to continue strengthening existing TB vaccine trial centres as well as establishing multiple settings, especially where TB prevalence is high. A next useful step will be the creation of a global directory of TB vaccine trial centres to reflect the need for TB vaccine development in geographies other than SSA.

## Data Availability

The data are publicly available on the EDCTP website: https://www.edctp.org/our-work/coordination-tb-vaccine-funded-research/directory-tb-vaccine-clinical-trial-sites-sub-saharan-africa/

## Acknowledgements

The directory of TB vaccine clinical trial centres suitable for future TB vaccine studies in Sub-Saharan Africa was commissioned and funded by the EDCTP2 programme supported by the European Union and conducted by TBVI and KNCV Tuberculosis foundation. The following experts provided advice on essential parameters that were included in the survey: Suzanne Verver, Frank Cobelens, Richard White and Rebecca Harris. We thank Ieva Leimane for her help with survey design, Daniela Pereira for data visualization, Degu Dare, Jeremy Hill, and Max Meis for technical support, and Stephanie Borsboom for project management. Our assessment relied partly on work done by KNCV for Aeras. Last but not least, we wish to thank all contact points from the centres for providing information.

## Author contributions

PTP conducted the data collection, developed the directory, summarized descriptions of the trial centres and wrote the paper. GV provided supervision and revisions to the paper. MH, PB, JVR, and TN provided revisions to the paper. MEHH and ALW commissioned the project and provided technical support and revisions to the paper. The funder was not involved in the analysis part.

---

**Supplement 2.** Overview of centres contacted, and centres responded per country by TB incidence, population size, and country size.

| Country | Population size | Country size (square Kilometre) | TB Incidence burden 2020 ( ) | Identified | Responded |
| --- | --- | --- | --- | --- | --- |
| Angola | 18,498,000 | 1,246,700 | 115 000 (74 000-165 000) |  |  |
| Benin | 8,791,832 | 112,62 | 6 700 (4 300-9 600) | 1 | 0 |
| Botswana | 1,990,876 | 600,37 | 5 600 (4 300-7 000) | 2 | 0 |
| Burkina Faso | 15,730,977 | 274,2 | 9 600 (6 200-14 000) | 1 | 1 |
| Burundi | 8,988,091 | 27,83 | 12 000 (8 000-18 000) |  |  |
| Cameroon | 18,879,301 | 475,44 | 46 000 (30 000-66 000) | 3 | 1 |
| Cape Verde | 499 | 322,462 | 220 (170-280) |  |  |
| Central African Republic | 4,511,488 | 622,984 | 26 000 (17 000-37 000) |  |  |
| Chad | 10,329,208 | 1,284,000 | 24 000 (15 000-34 000) |  |  |
| Comoros | 752,438 | 2,17 | 300 (200-430) |  |  |
| Democratic Republic of the Congo | 68,692,542 | 2,345,410 | 21 000 (13 000-30 000) | 1 | 0 |
| Djibouti | 516,055 | 23 | 2 200 (1 700-2 800) |  |  |
| Equatorial Guinea | 1,110,000 | 28,051 | 3 900 (2 500-5 600) |  |  |
| Eritrea | 5,647,168 | 121,32 | 2 900 (1 300-5 000) |  |  |
| Eswatini | 1,123,913 | 17,363 | 3 700 (2 300-5 400) | 1 | 1 |
| Ethiopia | 85,237,338 | 1,127,127 | 151 000 (106 000-205 000) | 3 | 2 |
| Gabon | 1,514,993 | 267,667 | 12 000 (7 600-17 000) | 1 | 0 |
| Gambia | 1,782,893 | 11,295 | 3 800 (2 800-4 900) | 2 | 0 |
| Ghana | 24,200,000 | 238,535 | 44 000 (21 000-76 000) | 3 | 1 |
| Guinea | 10,057,975 | 245,857 | 23 000 (15 000-34 000) |  |  |
| Guinea-Bissau | 1,647,000 | 36,125 | 7 100 (4 600-10 000) | 1 | 1 |
| Ivory Coast | 20,617,068 | 322,463 |  |  |  |
| Kenya | 39,002,772 | 582,65 | 139 000 (86 000-205 000) | 13 | 5 |
| Lesotho | 2,130,819 | 30,355 | 14 000 (8 600-20 000) |  |  |
| Liberia | 4,128,572 | 111,369 | 16 000 (10 000-23 000) |  |  |
| Madagascar | 19,625,000 | 587,041 | 66 000 (42 000-94 000) | 1 | 0 |
| Malawi | 14,268,711 | 118,48 | 27 000 (14 000-43 000) | 5 | 1 |
| Mali | 12,666,987 | 1,240,000 | 11 000 (6 900-15 000) | 1 | 1 |
| Mauritania | 3,359,185 | 1,030,700 | 4 000 (2 600-5 800) |  |  |
| Mauritius | 1,284,264 | 2,04 | 160 (120-200) |  |  |
| Mozambique | 21,669,278 | 801,59 | 115 000 (71 000-170 000) | 1 | 1 |
| Namibia | 2,108,665 | 825,418 | 12 000 (8 300-16 000) |  |  |
| Niger | 17,129,076 | 1,267,000 | 20 000 (13 000-29 000) |  |  |
| Nigeria | 174,507,539 | 923,768 | 452 000 (295 000-641 000) | 10 | 3 |
| Republic of the Congo | 3,700,000 | 342 | 286 000 (185 000-408 000) |  |  |
| Rwanda | 10,473,282 | 26,338 | 7 500 (5 700-9 600) | 2 | 0 |
| São Tomé and Príncipe | 212,679 | 1,001 | 260 (100-480) |  |  |
| Senegal | 12,855,153 | 196,712 | 20 000 (14 000-26 000) | 1 | 1 |
| Seychelles | 87,476 | 455 | 10 (9-12) |  |  |
| Sierra Leone | 6,190,280 | 71,74 | 24 000 (15 000-34 000) |  |  |
| Somalia | 9,832,017 | 637,657 | 41 000 (27 000-59 000) |  |  |
| South Africa | 59,899,991 | 1,219,912 | 328 000 (230 000-444 000) | 50 | 29 |
| South Sudan | 8,260,490 | 619,745 | 26 000 (17 000-37 000) |  |  |
| Sudan | 31,894,000 | 1,886,068 | 28 000 (19 000-38 000) |  |  |
| Tanzania | 44,928,923 | 945,087 | 133 000 (63 000-229 000) | 8 | 5 |
| Togo | 7,154,237 | 56,785 | 3 000 (2 400-3 600) |  |  |
| Uganda | 32,369,558 | 236,04 | 90 000 (54 000-135 000) | 13 | 2 |
| Zambia | 11,862,740 | 752,614 | 59 000 (38 000-84 000) | 6 | 1 |
| Zimbabwe | 11,392,629 | 390,58 | 29 000 (21 000-37 000) | 3 | 0 |

WHO TB incidence rates (13) / Worldbank population (14)

